# Epidemiology of zero-dose among children aged 12–35 months in Ethiopia: Burden, correlates and spatial patterns; Evidence from a 2025 national survey

**DOI:** 10.64898/2026.11.12.26367895

**Authors:** Mulugeta Geremew, Fisseha Shiferie, Asnake Worku, Abyot Asres, Ayalew Astatkie, Tigist Shumet, Dawit A. Tsegaye, Gashaw Andargie, Samson Gebremedhin

## Abstract

**Background:** Children who have not received the first dose of a diphtheria–tetanus–pertussis-containing vaccine are classified as zero-dose. Understanding how many children remain without this dose beyond infancy, the correlates, and their geographic distribution can inform routine and catch-up vaccination services in Ethiopia. This study aimed to estimate the burden of zero-dose, identify its correlates, and map its spatial distribution among children aged 12–35 months in Ethiopia.

**Methods:** We analyzed data from 2,097 children aged 12–35 months in the Pathways for Ethiopia survey, a recent national population-based cross-sectional household survey conducted from August to November 2025. The burden estimates and regression analyses accounted for sampling weights, stratification, and clustering. Adjusted prevalence ratios (aPRs) were obtained through marginal standardization of survey-weighted logistic regression models. Spatial autocorrelation, hotspot, and spatial-scan analyses assessed geographic clustering. Ordinary kriging predicted prevalence at unsampled locations.

**Results:** The weighted prevalence of zero-dose status was 20.6% (95% confidence interval [CI]: 15.9–26.3), with regional point estimates ranging from 0.8% in Addis Ababa to 35.9% in Somali. Its burden was 20.4% among children aged 12–23 months and 20.8% among those aged 24–35 months. Higher burden was associated with no antenatal care compared with four or more visits (aPR: 2.39; 95% CI: 1.61–3.53), unimproved compared with improved drinking-water sources (aPR: 1.52; 95% CI: 1.18–1.97), and caregivers rating vaccination as less than very important compared with very important (aPR: 2.68; 95% CI: 2.03–3.54). Zero-dose prevalence showed significant spatial clustering (Moran’s I = 0.229; p = 0.0001). Hotspots occurred among sampled communities in Somali, Oromia, Central Ethiopia, Afar, and South Ethiopia. Predicted prevalence was highest across parts of eastern and south-central Ethiopia.

**Conclusion:** Approximately one in five children aged 12–35 months had not received the first pentavalent dose. High zero-dose prevalence clustered among sampled communities in eastern and south-central Ethiopia. Moreover, antenatal care and drinking water source were independent factors. Immunization programs should prioritize these communities for identifying and vaccinating missed children, including those aged 24–35 months.

## Background

Childhood immunization prevents serious illness and death, yet millions of children continue to miss essential vaccine doses. Children who have not received the first dose of a diphtheria– tetanus–pertussis-containing vaccine (DTP1) are classified as zero-dose [1]. Globally, an estimated 13.5 million children were zero-dose in 2025 [2]. Missing this first dose is an important indicator of gaps in the reach of routine immunization services. Reaching these children is central to the Immunization Agenda 2030 commitment to equitable access to vaccination [1].

Ethiopia has a substantial burden of zero-dose children, with considerable geographic variation. Analyses of the 2019 Mini Demographic and Health Survey and the 2023 immunization survey estimated zero-dose prevalence at 23.7% and 24.8%, respectively, among children aged 12–23 months, which indicated that its prevalence is high in Somali and much lower prevalence in Addis Ababa [3,4]. Local concentrations of children who missed the first dose have also been identified in Somali, Afar, and parts of Amhara and Oromia [5]. These patterns show that the national burden is unevenly distributed and that regional averages may not show local concentrations of high zero-dose prevalence. Research among children aged 12–35 months in selected underserved settings similarly documented considerable local differences [5].

Fewer antenatal care visits, delivery outside health institutions, and rural residence were identified as factors associated with zero-dose from the finding generated in the 2019 survey [3]. Consistently, finding generated from 2023 survey showed that antenatal care, perceived vaccination benefits, and distance to vaccination services, with differences in these relationships across locations were factors associated with zero-dose[4]. These findings identify maternal healthcare contact and vaccination perceptions as relevant factors for further investigation [3,4]. Ethiopia introduced a catch-up vaccination policy in 2022 and renewed implementation of the Big Catch-Up initiative in 2025 [6]. To minimize the zero dose status among children, routine immunization services were strengthened, underserved and hard to reach populations were reached, community engagement and health education, strengthening health extension programs and integration of immunization with other essential health services were implemented as strategies in Ethiopia [6].

The 2023 national spatial analysis also excluded Tigray because of security constraints [4]. These studies establish the importance of geographic and population differences, while leaving a need for updated national evidence that includes older children who remain without a first dose. As catch-up services expand, understanding the size and geographic distribution of this unmet vaccination need can help determine whom to reach and where to focus vaccination activities.

Using a national population-based household survey conducted in 2025, this study aimed to estimate national and regional zero-dose prevalence among children aged 12–35 months in Ethiopia, identify correlates among child, maternal, household, and service characteristics, and examine spatial clustering and geographic patterns of zero-dose status.

## Methods

### Study design, setting, and participants

This study used data from the Pathways for Ethiopia national population-based cross-sectional household survey, conducted from August to November 2025 across all 12 regional states and the two chartered cities. Information on children was collected in the survey of women of reproductive age. The survey used a stratified two-stage cluster sampling design. In the first stage, census enumeration areas (EAs) were stratified by region and urban–rural residence and selected for the survey. In the second stage, eligible women were listed and sampled within the selected EAs. Fieldwork was completed in 247 of the 251 planned EAs. The analysis included children aged 12–35 months whose first pentavalent dose status could be determined from the survey. Records with incomplete or invalid sampling or geographic information were excluded.

### Variables and measurements

Zero-dose status was defined as nonreceipt of the first pentavalent dose, a diphtheria–tetanus– pertussis-containing vaccine. Vaccination status was assessed from vaccination records and caregiver reports, with records taking precedence when available. The independent variables were child age, region, residence, maternal age, maternal education, household financial situation, water and sanitation, antenatal care (ANC), place of delivery, walking time to the nearest health facility, self-reported local armed conflict, participation in decisions about child healthcare, and perceived vaccination importance. Caregivers rated the importance of vaccines for their child’s health on a four-point scale: “not at all important,” “a little important,” “moderately important,” or “very important” [7]. For analysis, the first three responses were grouped as “less than very important,” with “very important” as the reference category.

### Survey-weighted analysis

Data were analyzed using R version 4.3.3 [8]. The national and regional zero dose prevalence was estimated using the final normalized survey weights, accounting for clustering within 247 EAs and stratification by region and urban–rural residence (27 strata). Design-based 95% confidence intervals were calculated on the logit scale and back-transformed. Regional estimates with coefficients of variation of 0.30 or greater were considered imprecise.

Survey-weighted logistic regression was used to examine associations with zero-dose status. The adjusted analysis included 2,015 children with complete data for all model variables. Adjusted prevalence ratios (aPRs) and 95% confidence intervals were obtained by marginal standardization of predicted probabilities, adjusting for region, residence and the other covariates. Overall design-based Wald tests assessed factors with more than two categories.

### Spatial analysis

Spatial autocorrelation in zero-dose prevalence was assessed using global Moran’s I. Local Moran’s I was used to identify local clusters and spatial outliers, while Getis–Ord Gi* identified hotspots and cold spots [9,10]. Local p values were adjusted for multiple comparisons using the Benjamini–Hochberg false discovery rate procedure. A Bernoulli spatial scan was used to identify geographic clusters of high zero-dose prevalence, with zero-dose children classified as cases and other eligible children as controls [11]. The primary cluster and nonoverlapping secondary clusters were mapped.

Ordinary kriging was used to predict zero-dose prevalence at unsampled locations using survey-weighted estimates from sampled EAs [4]. Linear, spherical and exponential semivariograms were compared using leave-one-EA-out cross-validation. Predictions were generated on a 10-km grid using the 16 nearest EAs. The exponential model was further evaluated using five geographically separated validation groups. Its prediction error was compared with that of a prediction based on the pooled prevalence in the training sample.

Geographic variation in associations with zero-dose status was examined using binomial models with a spatial intercept and a spatially varying coefficient. Ten contrasts involving child, maternal, household and service characteristics were assessed among children with complete model data. Tests of geographic variation were adjusted for multiple comparisons, and spatial cross-validation compared each model with a corresponding constant-coefficient model. Local odds ratios were mapped for contrasts showing geographic variation and improved prediction, at locations with sufficient observations in both comparison groups. Each odds ratio compared the specified category with its reference group.

## Results

### The burden of zero-dose among children

The study included 2,097 children aged 12–35 months in 247 sampled EAs, of whom 311 were zero-dose. National survey-weighted prevalence of zero-dose was 20.6% (95% CI 15.9–26.3). The estimate varied across regions, from 35.9% (17.4–59.9) in Somali and 32.1% (22.2–43.8) in Oromia to 0.8% (0.1–5.5) in Addis Ababa (Fig 1). This burden varied as 20.4% among children aged 12–23 months and 20.8% among those aged 24–35 months (Table 1).

**Fig 1.**
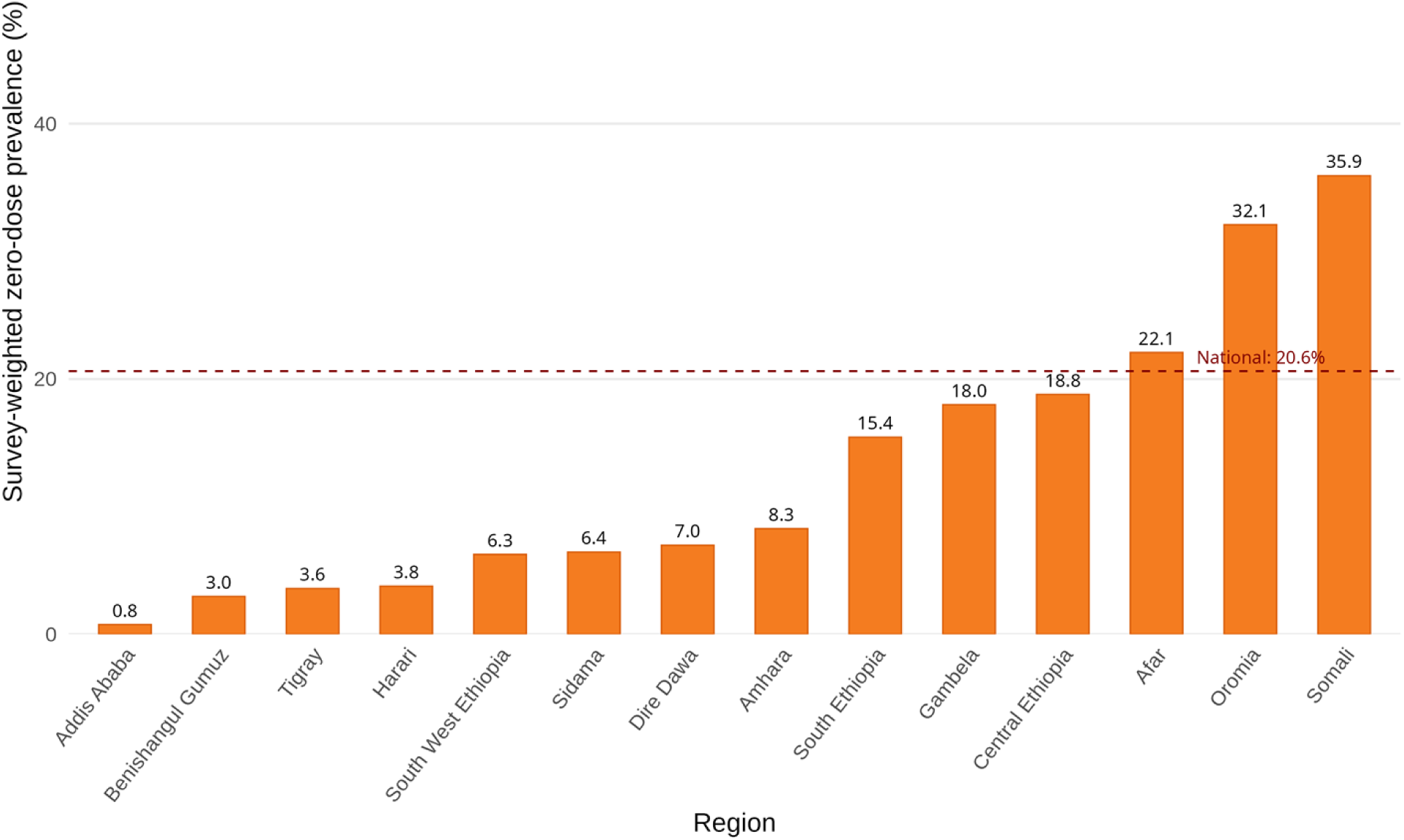
Survey-weighted prevalence of zero-dose children aged 12–35 months by region, Ethiopia, 2025.

**Table 1.**
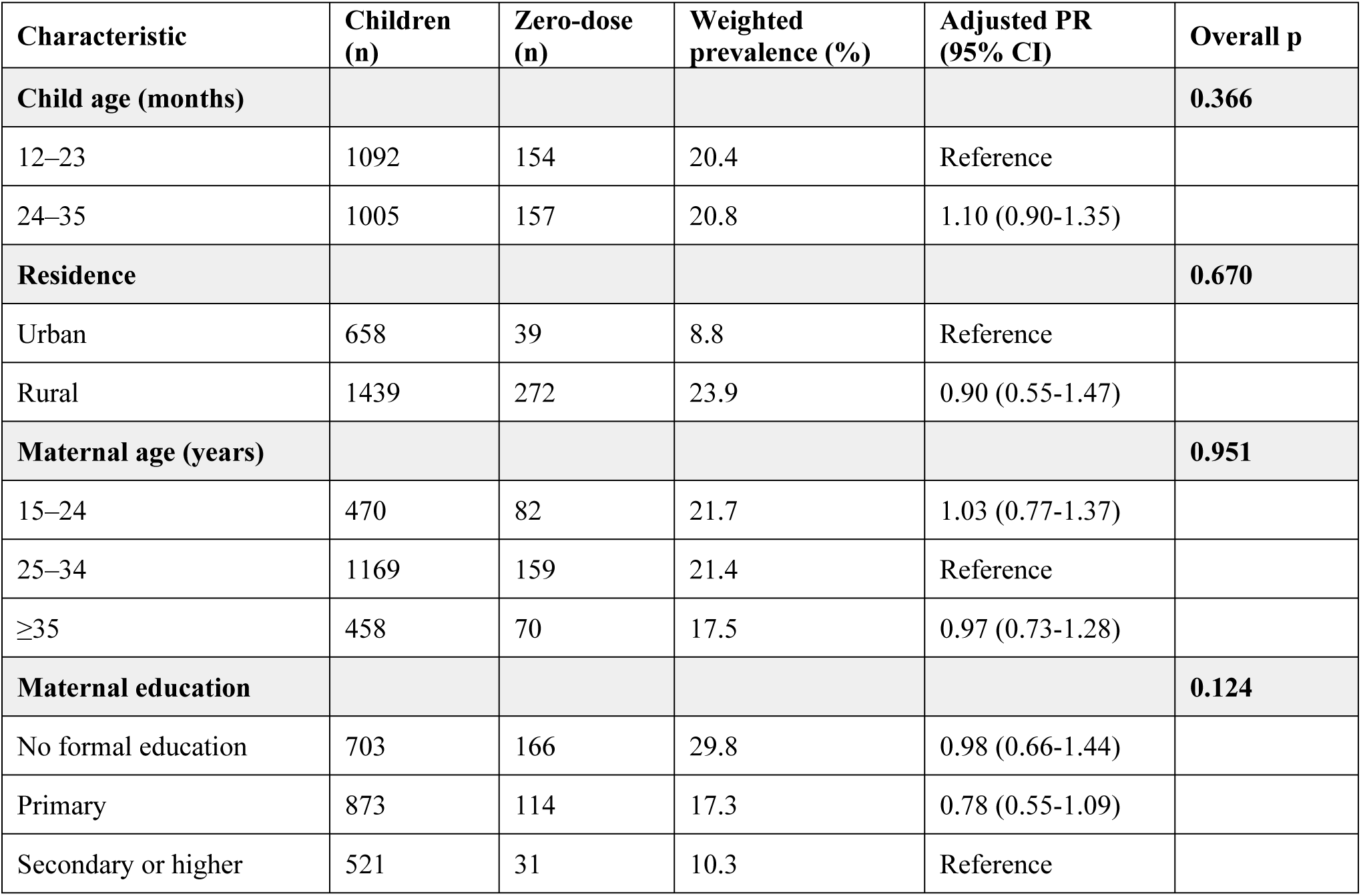
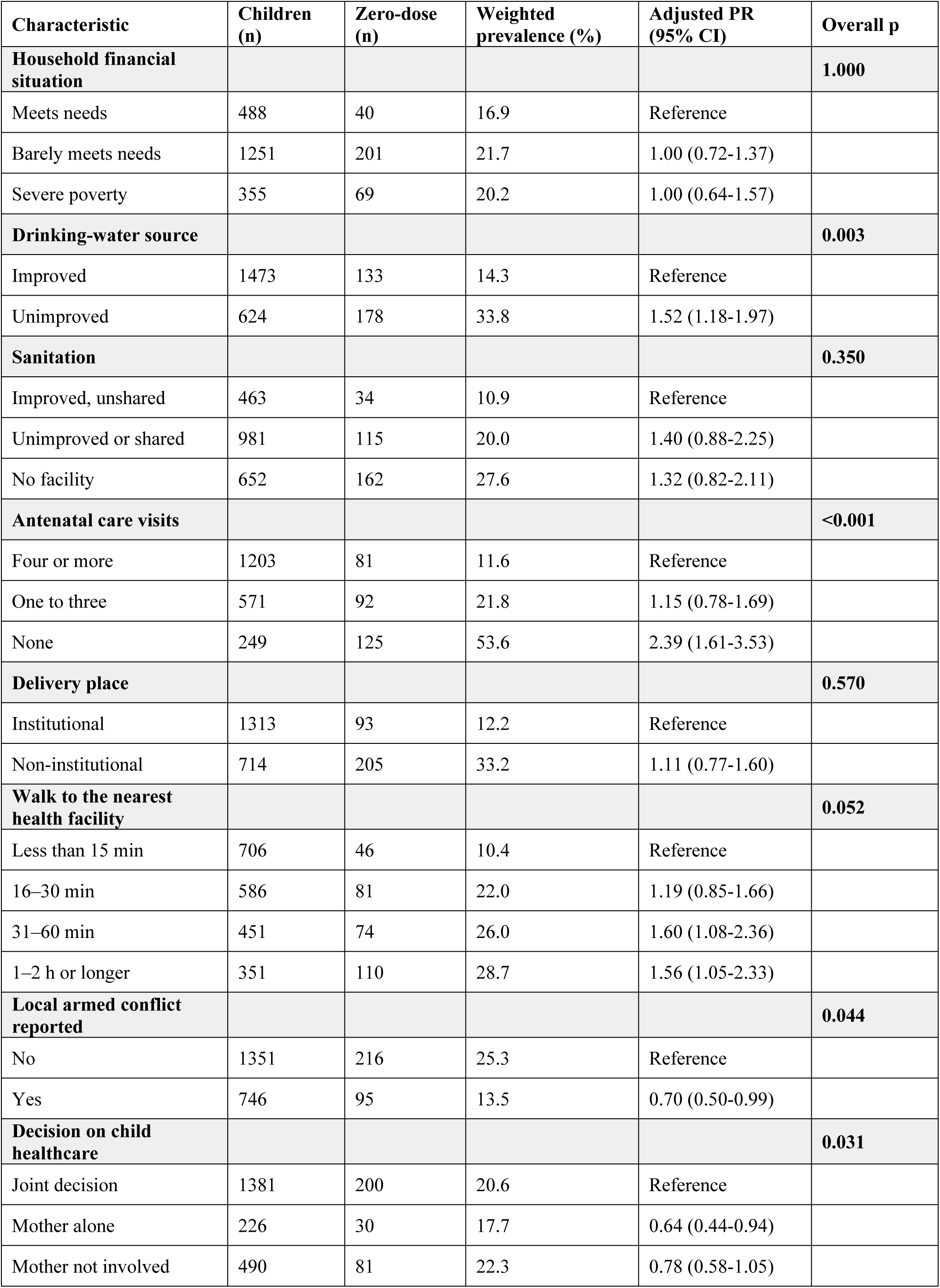
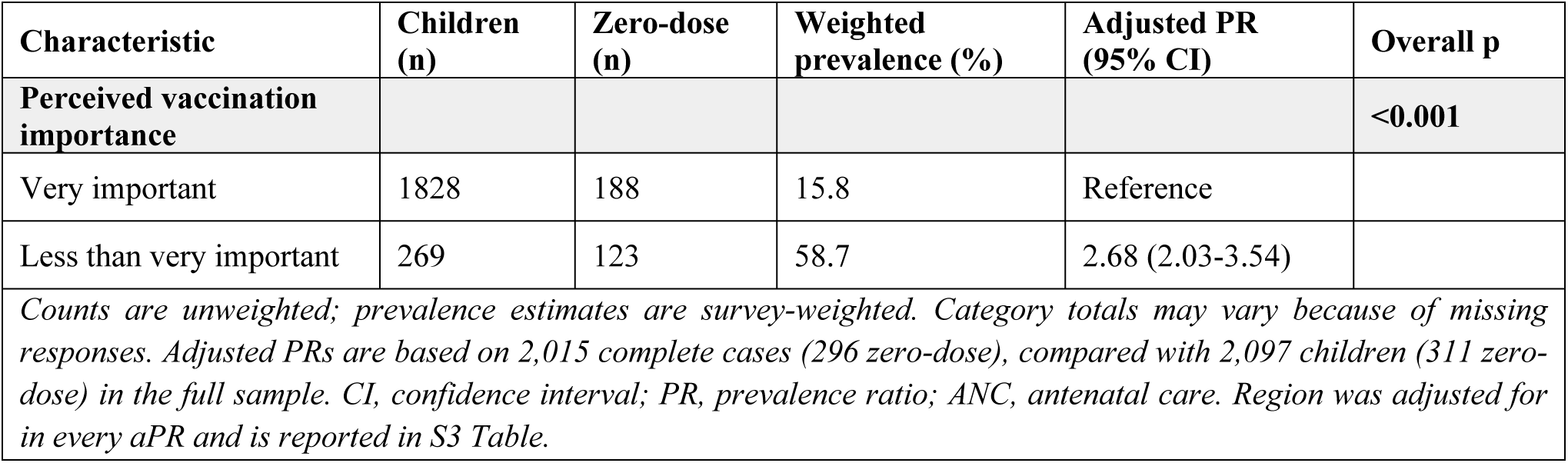
Zero-dose prevalence and correlates among children aged 12–35 months, Ethiopia, 2025.

| **Characteristic** | **Children (n)** | **Zero-dose (n)** | **Weighted prevalence (%)** | **Adjusted PR (95% CI)** | **Overall p** |
|---|---|---|---|---|---|
| **Child age (months)** |  |  |  |  | **0.366** |
| 12–23 | 1092 | 154 | 20.4 | Reference |  |
| 24–35 | 1005 | 157 | 20.8 | 1.10 (0.90-1.35) |  |
| **Residence** |  |  |  |  | **0.670** |
| Urban | 658 | 39 | 8.8 | Reference |  |
| Rural | 1439 | 272 | 23.9 | 0.90 (0.55-1.47) |  |
| **Maternal age (years)** |  |  |  |  | **0.951** |
| 15–24 | 470 | 82 | 21.7 | 1.03 (0.77-1.37) |  |
| 25–34 | 1169 | 159 | 21.4 | Reference |  |
| ≥35 | 458 | 70 | 17.5 | 0.97 (0.73-1.28) |  |
| **Maternal education** |  |  |  |  | **0.124** |
| No formal education | 703 | 166 | 29.8 | 0.98 (0.66-1.44) |  |
| Primary | 873 | 114 | 17.3 | 0.78 (0.55-1.09) |  |
| Secondary or higher | 521 | 31 | 10.3 | Reference |  |
| **Characteristic** | **Children (n)** | **Zero-dose (n)** | **Weighted prevalence (%)** | **Adjusted PR (95% CI)** | **Overall p** |
| **Household financial situation** |  |  |  |  | **1.000** |
| Meets needs | 488 | 40 | 16.9 | Reference |  |
| Barely meets needs | 1251 | 201 | 21.7 | 1.00 (0.72-1.37) |  |
| Severe poverty | 355 | 69 | 20.2 | 1.00 (0.64-1.57) |  |
| **Drinking-water source** |  |  |  |  | **0.003** |
| Improved | 1473 | 133 | 14.3 | Reference |  |
| Unimproved | 624 | 178 | 33.8 | 1.52 (1.18-1.97) |  |
| **Sanitation** |  |  |  |  | **0.350** |
| Improved, unshared | 463 | 34 | 10.9 | Reference |  |
| Unimproved or shared | 981 | 115 | 20.0 | 1.40 (0.88-2.25) |  |
| No facility | 652 | 162 | 27.6 | 1.32 (0.82-2.11) |  |
| **Antenatal care visits** |  |  |  |  | **<0.001** |
| Four or more | 1203 | 81 | 11.6 | Reference |  |
| One to three | 571 | 92 | 21.8 | 1.15 (0.78-1.69) |  |
| None | 249 | 125 | 53.6 | 2.39 (1.61-3.53) |  |
| **Delivery place** |  |  |  |  | **0.570** |
| Institutional | 1313 | 93 | 12.2 | Reference |  |
| Non-institutional | 714 | 205 | 33.2 | 1.11 (0.77-1.60) |  |
| **Walk to the nearest health facility** |  |  |  |  | **0.052** |
| Less than 15 min | 706 | 46 | 10.4 | Reference |  |
| 16–30 min | 586 | 81 | 22.0 | 1.19 (0.85-1.66) |  |
| 31–60 min | 451 | 74 | 26.0 | 1.60 (1.08-2.36) |  |
| 1–2 h or longer | 351 | 110 | 28.7 | 1.56 (1.05-2.33) |  |
| **Local armed conflict reported** |  |  |  |  | **0.044** |
| No | 1351 | 216 | 25.3 | Reference |  |
| Yes | 746 | 95 | 13.5 | 0.70 (0.50-0.99) |  |
| **Decision on child healthcare** |  |  |  |  | **0.031** |
| Joint decision | 1381 | 200 | 20.6 | Reference |  |
| Mother alone | 226 | 30 | 17.7 | 0.64 (0.44-0.94) |  |
| Mother not involved | 490 | 81 | 22.3 | 0.78 (0.58-1.05) |  |
| **Characteristic** | **Children (n)** | **Zero-dose (n)** | **Weighted prevalence (%)** | **Adjusted PR (95% CI)** | **Overall p** |
| **Perceived vaccination importance** |  |  |  |  | **<0.001** |
| Very important | 1828 | 188 | 15.8 | Reference |  |
| Less than very important | 269 | 123 | 58.7 | 2.68 (2.03-3.54) |  |

*Counts are unweighted; prevalence estimates are survey-weighted. Category totals may vary because of missing responses. Adjusted PRs are based on 2,015 complete cases (296 zero-dose), compared with 2,097 children (311 zero-dose) in the full sample. CI, confidence interval; PR, prevalence ratio; ANC, antenatal care. Region was adjusted for in every aPR and is reported in S3 Table.*

### Correlates of zero-dose

The adjusted model included 2,015 children in 247 sampled EAs, of whom 296 were zero-dose (Table 1). Children whose mothers had not attended any ANC visits had higher zero-dose prevalence than children whose mothers reported four or more visits (aPR: 2.39, 95% CI: 1.61–3.53). Children living in households with unimproved drinking water sources had 52 percent higher prevalence of zero-dose as compared with those living in households with improved drinking water sources (aPR: 1.52, 95% CI: 1.18–1.97). Consistently, children whose caregivers perceived vaccination as less than very important had 2.68 times the prevalence of zero-dose as compared with children whose caregivers considered vaccination very important (aPR: 2.68, 95% CI: 2.03–3.54).

For walking time to the nearest health facility, 31–60 minutes (aPR: 1.60, 95% CI: 1.08–2.36) and 1–2 hours or longer (aPR: 1.56, 95% CI: 1.05–2.33) were associated with higher prevalence than less than 15 minutes, although the overall association with walking time was not statistically significant (p = 0.052). Reported local armed conflict (aPR: 0.70, 95% CI: 0.50–0.99) and mothers deciding alone rather than jointly on child healthcare (aPR: 0.64, 95% CI: 0.44–0.94) were associated with lower prevalence. Child age, maternal age, residence, maternal education, household financial situation, sanitation and delivery place had no statistically significant overall adjusted associations. Adjusted regional contrasts are presented in S3 Table.

### Geographic patterns of zero-dose children

Zero-dose prevalence was geographically clustered, with neighbouring sampled communities tending to have similar prevalence (Moran’s I = 0.229; p = 0.0001; Table 2; Fig 2).

**Table 2.** Global and local spatial analysis of zero-dose prevalence among children aged 12– 35 months, Ethiopia, 2025.

| Analysis | Estimate | Inference | Interpretation |
|---|---|---|---|
| Global Moran’s I; 8 nearest neighbours | 0.229 | 9,999 unrestricted permutations; p=0.0001 | Positive spatial association |
| Local Moran’s I; FDR-adjusted | 11 HH; 6 LH | Benjamini–Hochberg q<0.05 | High–high clusters and low–high outliers |
| Getis-Ord Gi*; FDR-adjusted | 25 high; 0 low | Benjamini-Hochberg q<0.05 | High-value concentrations in five regions |

Global Moran’s I was calculated from survey-weighted prevalence at sampled EAs; local classifications use Benjamini–Hochberg-adjusted q < 0.05.

**Fig 2.**
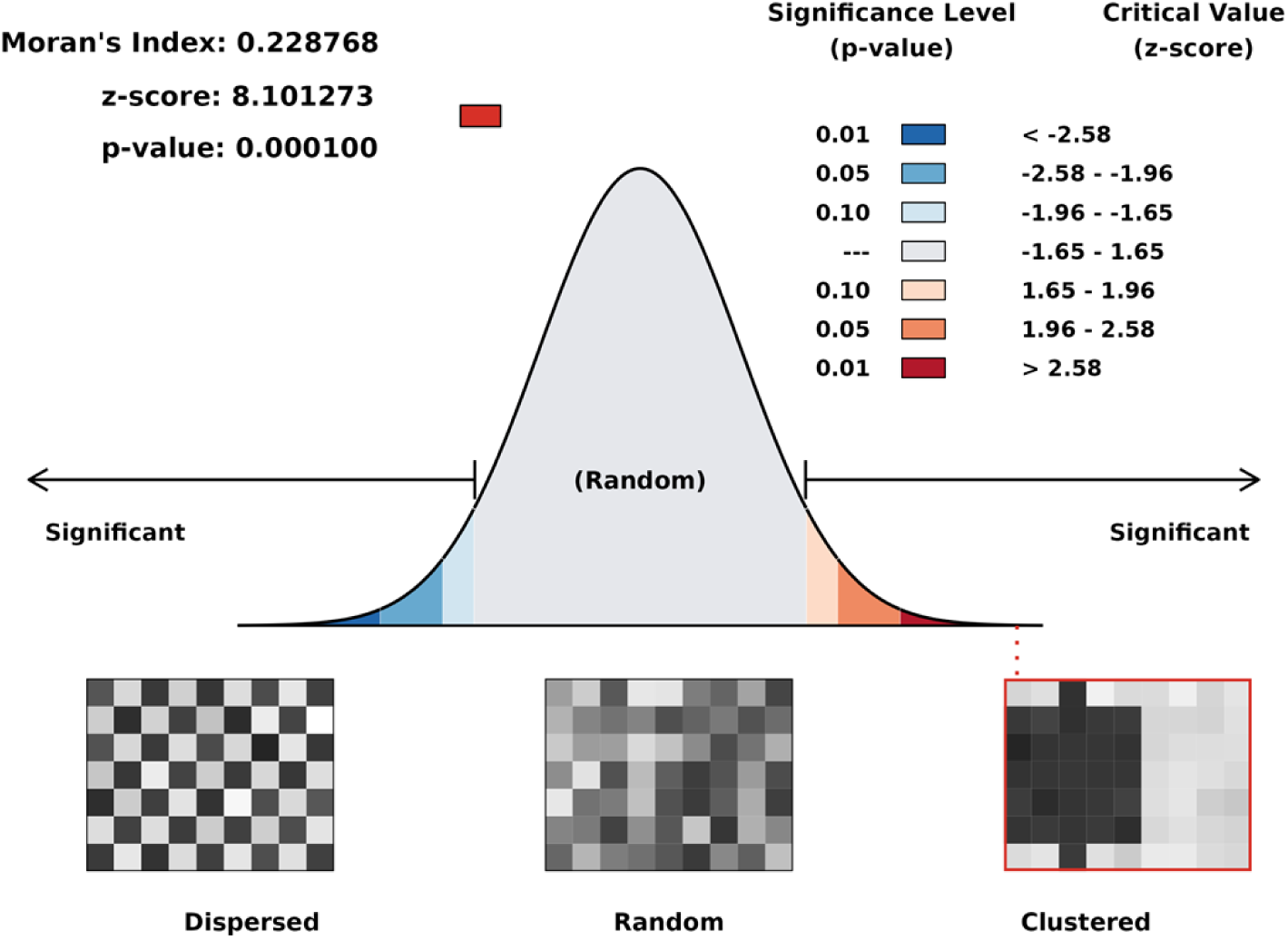
Spatial autocorrelation of zero-dose prevalence among children aged 12–35 months, Ethiopia, 2025. Local Moran’s I identified 11 sampled communities with high zero-dose prevalence surrounded by communities with similarly high prevalence (high–high clusters; q < 0.05; Fig 3; S2 Table). These were in Somali (four), Oromia (three), Central Ethiopia (two), Afar (one) and South Ethiopia (one). Six communities with low prevalence were surrounded by communities with high prevalence (low–high outliers): four in Somali, one in Afar and one in South Ethiopia.

**Fig 3.**
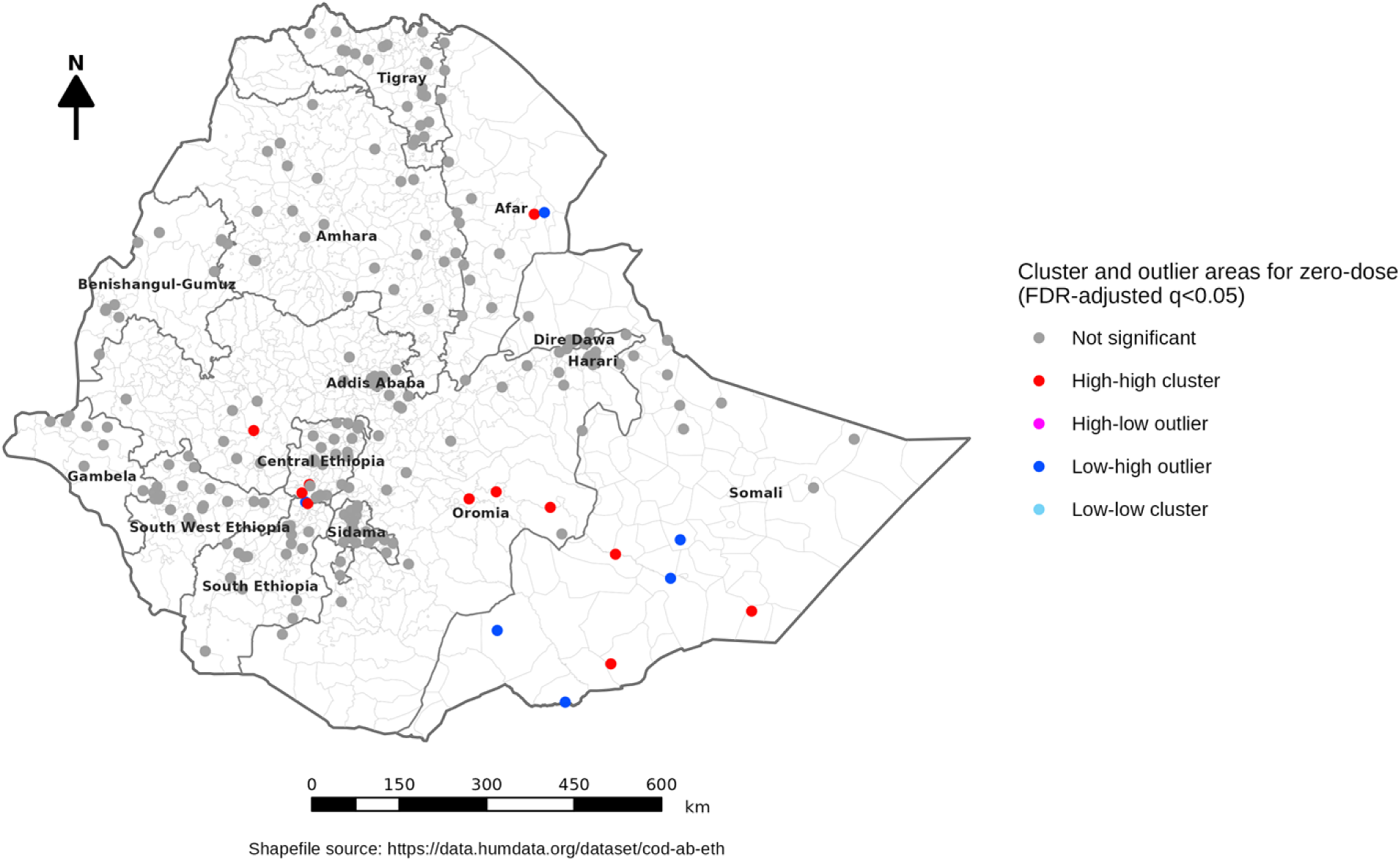
Local Moran clusters and outliers of zero-dose prevalence among sampled communities, Ethiopia, 2025. EA, enumeration area; FDR, false discovery rate. Local significance was assessed at Benjamini–Hochberg-adjusted q < 0.05.

Hotspots were concentrated in Somali, Oromia and Central Ethiopia, with additional hotspots in Afar and South Ethiopia (Fig 4). Getis–Ord Gi* identified 25 hotspot communities at q < 0.05: nine in Somali, six in Oromia, six in Central Ethiopia, two in Afar and two in South Ethiopia. Eleven met q < 0.01. No cold spots met q < 0.05.

**Fig 4.**
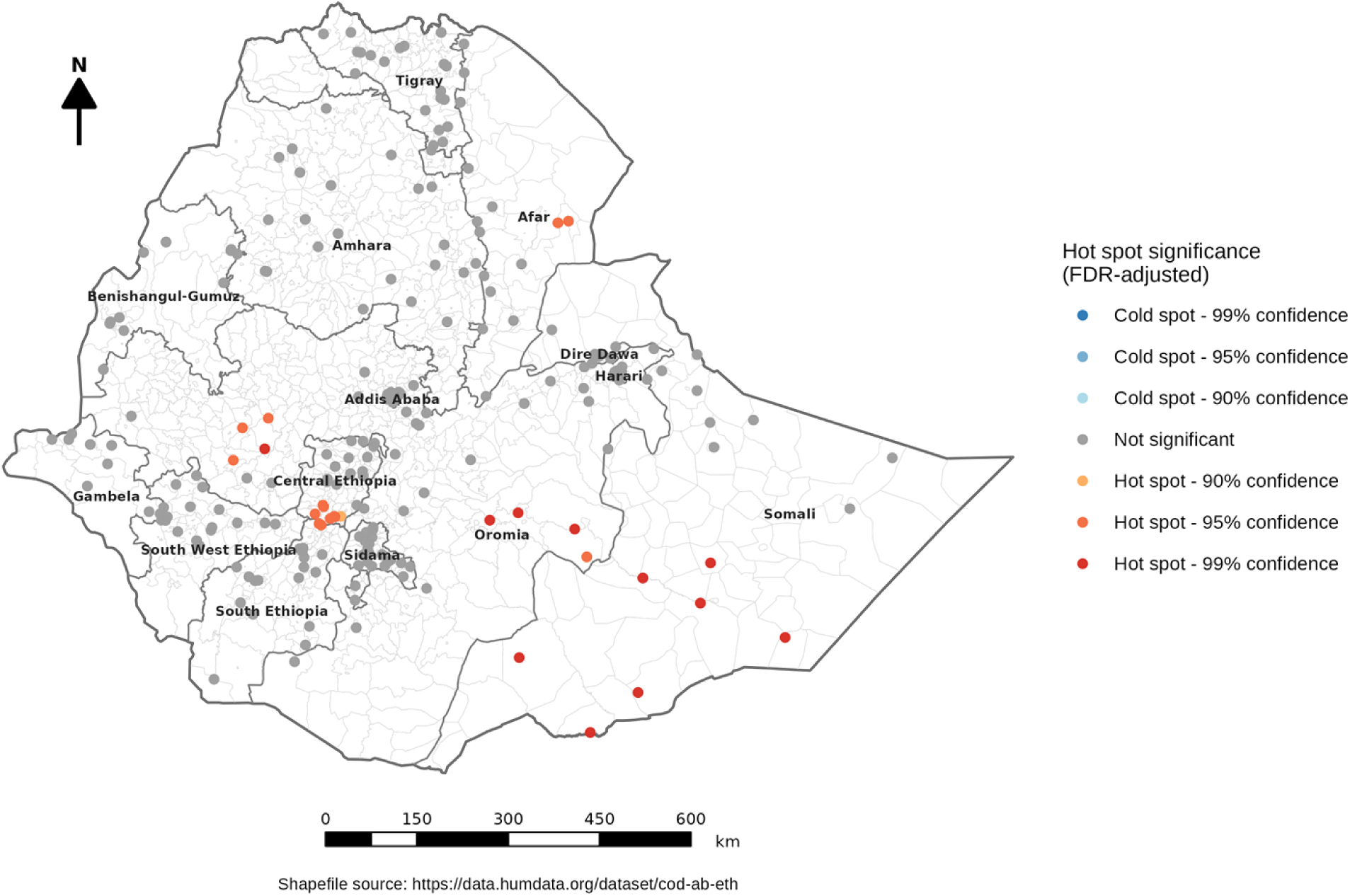
Hotspots and cold spots of zero-dose prevalence among sampled communities, Ethiopia, 2025. FDR, false discovery rate. Classes correspond to adjusted q < 0.10, q < 0.05, and q < 0.01.

The spatial scan identified one primary and six secondary clusters of high zero-dose prevalence (Table 3; Fig 5). The primary cluster extended across parts of Somali, Oromia and Sidama and included 58 zero-dose children among 109 sampled children in 11 communities (relative risk 4.18; p = 0.001). Three secondary clusters also had p = 0.001: Oromia (30 of 60 children), Afar– Amhara (41 of 107) and Central–South Ethiopia (31 of 77).

**Table 3.** Primary and secondary Bernoulli spatial-scan clusters of zero-dose children aged 12–35 months, Ethiopia, 2025.

| **Rank** | **Type** | **Survey EAs** | **Children** | **Zero-dose** | **LLR** | **RR** | **P** | **Radius in km** |
|---|---|---|---|---|---|---|---|---|
| 1 | Primary | 11 | 109 | 58 | 47.18 | 4.18 | 0.001 | 241.5 |
| 2 | Secondary | 6 | 60 | 30 | 21.36 | 3.62 | 0.001 | 102.3 |
| 3 | Secondary | 9 | 107 | 41 | 18.91 | 2.82 | 0.001 | 158.1 |
| 4 | Secondary | 10 | 77 | 31 | 15.41 | 2.90 | 0.001 | 48.0 |
| 5 | Secondary | 6 | 60 | 22 | 8.99 | 2.58 | 0.022 | 109.5 |
| 6 | Secondary | 1 | 12 | 8 | 8.35 | 4.59 | 0.039 | 0.0 |
| 7 | Secondary | 2 | 15 | 9 | 8.13 | 4.14 | 0.043 | 44.9 |

LLR, log-likelihood ratio; RR, relative risk. Clusters refer to the sampled communities within each scan window.

**Fig 5.**
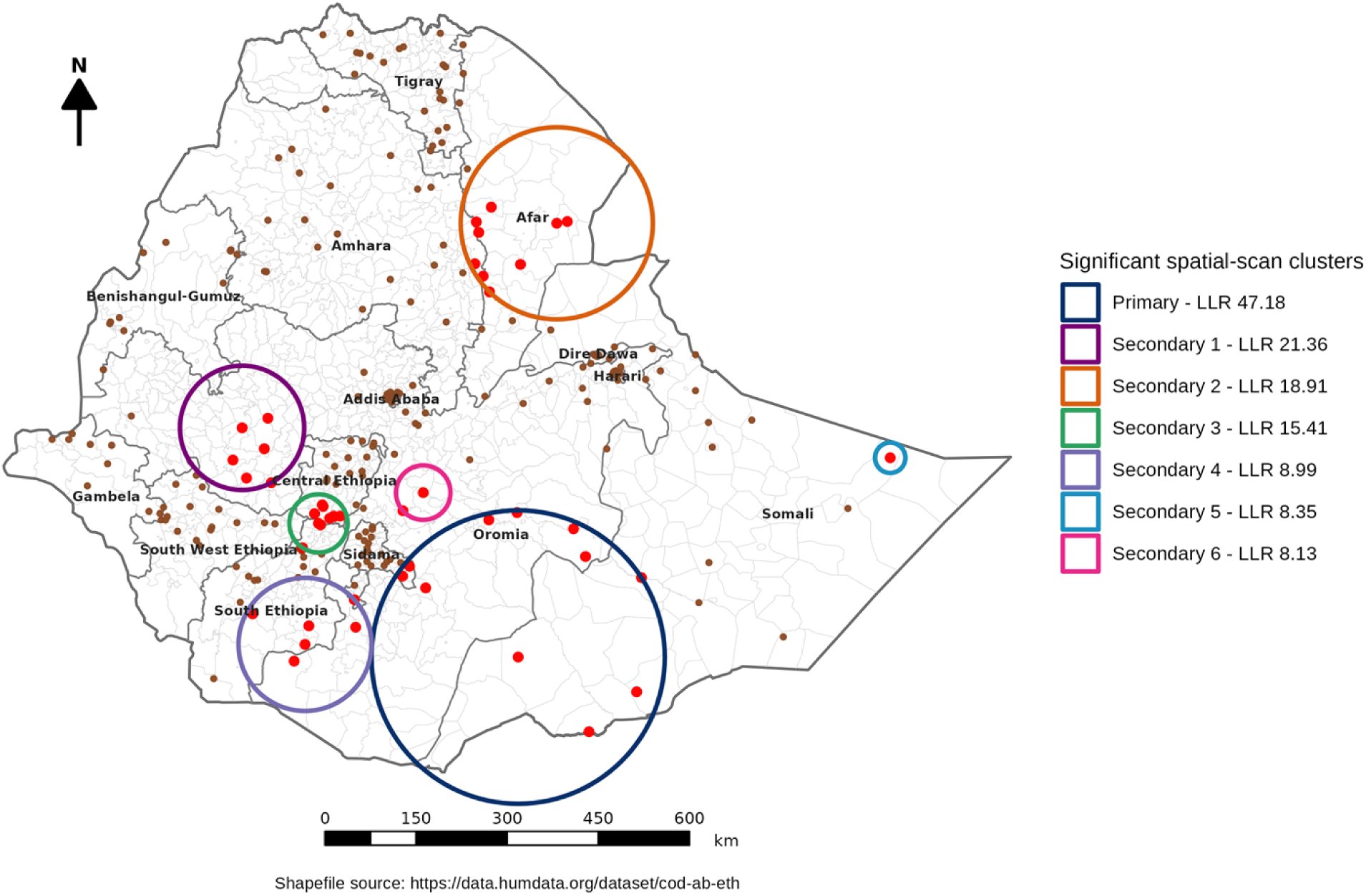
Spatial-scan clusters of zero-dose children among sampled communities, Ethiopia, 2025. LLR, log-likelihood ratio. Rings identify the primary and six secondary scan windows.

### Predicted prevalence of zero-dose children

The predicted surface showed higher zero-dose prevalence across parts of eastern and south-central Ethiopia (Fig 6). The exponential semivariogram had the smallest leave-one-EA-out prediction error (RMSE 19.44 percentage points; Table 4). In five geographically separated validation folds, its RMSE was 21.66 percentage points, compared with 23.00 for predictions based on the pooled prevalence in the training sample.

**Table 4.** Leave-one-EA-out prediction error for candidate ordinary-kriging models.

| **Semivariogram** | **EAs** | **ME** | **MAE** | **RMSE** | **Selection** |
|---|---|---|---|---|---|
| Exponential | 247 | 1.99 | 13.52 | 19.44 | Selected |
| Spherical | 247 | 2.21 | 13.59 | 19.59 | Candidate |
| Linear | 247 | 2.28 | 13.59 | 19.61 | Candidate |

Prediction errors are percentage points. ME, mean error; MAE, mean absolute error; RMSE, root mean squared error.

**Fig 6.**
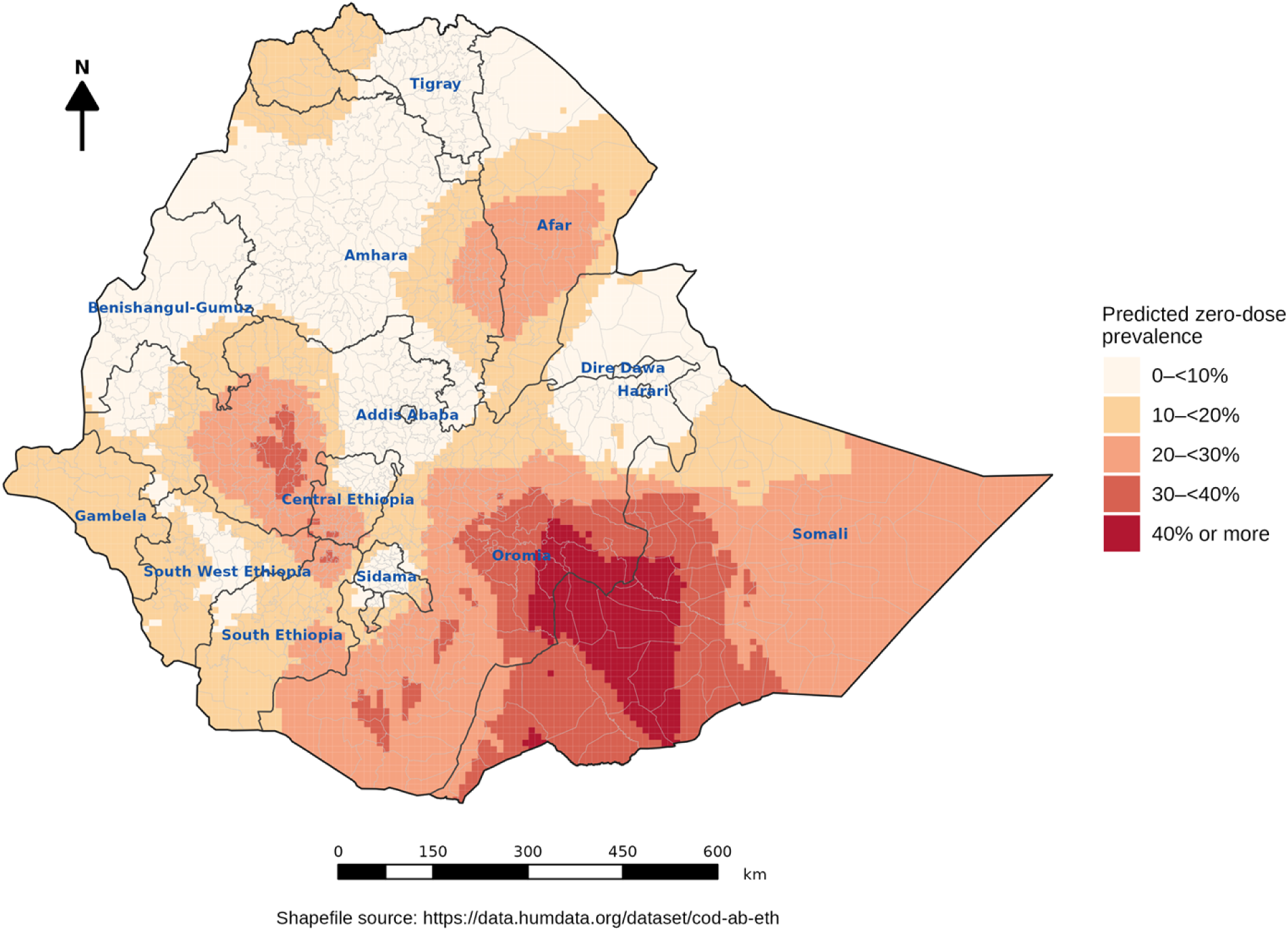
Predicted zero-dose prevalence among children aged 12–35 months, Ethiopia, 2025. Ordinary kriging estimates on a 10-km grid.

### Geographic variation in local associations

The associations of ANC attendance and participation in decisions about child healthcare with zero-dose status varied across sampled communities (Fig 7; S4 Table). For one to three compared with at least four ANC visits (q = 0.015), local odds ratios ranged from 0.23 to 2.09 across 105 communities with adequate local data, with higher values in some central sampled communities. For no participation compared with any participation in decisions about child healthcare (q = 0.006), local odds ratios ranged from 0.15 to 1.97 across 123 communities. Higher values occurred in several northern and eastern sampled communities, whereas lower values occurred in some southwestern communities. Both spatially varying models performed slightly better than the constant-coefficient models in spatial cross-validation.

**Fig 7.**
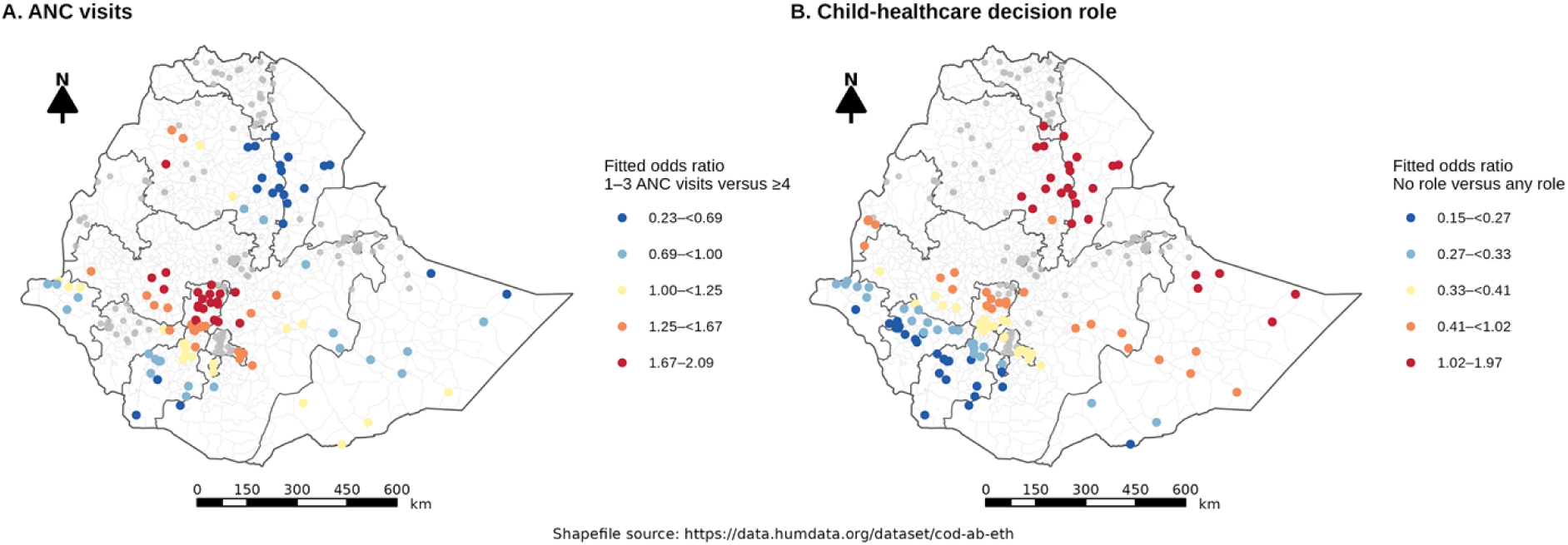
Geographic variation in odds ratios for antenatal care and participation in decisions about child healthcare, Ethiopia, 2025. Panel A compares 1–3 with ≥4 antenatal care (ANC) visits; panel B compares no participation with any participation in decisions about child healthcare. Grey points indicate sampled communities without adequate local comparison-group data.

## Discussion

Approximately one in five children aged 12–35 months had not received the first pentavalent dose in 2025. Zero-dose status was associated with maternal healthcare use, household living conditions and perceptions of vaccination. High zero-dose prevalence clustered in sampled communities in eastern and south-central Ethiopia. These findings show that missed first doses remain a substantial concern beyond infancy and identify communities where vaccination services should be strengthened.

The 20.6% prevalence (95% CI: 15.9–26.3) was broadly comparable to the 23.7% reported by Muchie and colleagues from the 2019 Mini Demographic and Health Survey of children aged 12–23 months [3] and the 24.8% reported by Endehabtu and colleagues from a 2023 programme evaluation of the same younger age group in ten regions and two cities [4]. Tamir and Agimas also reported geographic variation using other spatial analyses of the 2019 survey [12,13]. Selected underserved Ethiopian communities had approximately one-third of children classified as zero-dose [5,14]. Differences in age groups, sampled populations and zero-dose definitions may contribute to the variation between estimates and limit conclusions about changes over time [15].

Somali and Oromia had the highest regional point estimates in this survey, although regional confidence intervals were often wide. Endehabtu and colleagues likewise reported elevated prevalence in Somali and Afar, while their 2023 survey could not include Tigray; Shiferie and colleagues identified concentrations in Somali and Afar in underserved settings [4,5]. Similar geographic differences have been reported for broader childhood vaccination outcomes in Ethiopia [16–20]. Spatial studies of BCG and measles vaccination have also documented geographic differences within Ethiopia [21–23]. The repeated identification of high-prevalence areas supports prioritizing these communities when planning outreach and identifying children who have missed vaccination.

ANC was found to be a factor associated with zero-dose among children. Children whose mothers reported no ANC visits had higher zero-dose prevalence as compared with those mothers reported four or more visits. This finding is consistent with the study conducted in Kenya [24] and Ethiopia in 2019 [3]. Interviews in underserved Ethiopian communities documented irregular immunization sessions and inadequate information from health workers [25]. The observed association between ANC attendance and childhood vaccination may also reflect broader engagement with the healthcare system, as mothers who attend ANC may be more likely to use immunization services for their children. ANC visits may provide opportunities for mothers to receive information about childhood vaccination schedules and develop trust in vaccination services [4].

Lower perceived vaccination importance was also strongly associated with zero-dose status. Endehabtu and colleagues similarly reported an association involving caregivers’ perceptions of vaccination benefits [4]. Reviews of childhood immunization have identified parental attitudes, limited information and communication barriers among the reasons for non-vaccination [26,27]. These findings support clear explanations of vaccination benefits and opportunities for caregivers to discuss their concerns. Because perceptions and vaccination status were measured at the same time, present views may precede missed vaccination or reflect previous service encounters.

An unimproved drinking-water source was noted as a factor associated with zero-dose status. Children living in households with unimproved drinking water sources had 52% higher prevalence of zero-dose as compared with those living in households with improved drinking water sources. Water source may reflect household living conditions and access to basic services. Wendt and colleagues, using surveys from 80 low- and middle-income countries, found that children who had received no DTP-containing vaccine were more likely to experience other forms of deprivation, with particularly consistent associations involving water, sanitation and maternal education [28]. An analysis across 33 sub-Saharan African countries also linked receipt of no vaccine with household wealth and maternal healthcare use [29]. Limited access to improved water and vaccination may therefore reflect overlapping household disadvantages. The finding supports considering broader living conditions when identifying children missed by immunization services.

In the descriptive analysis, zero-dose prevalence was higher among rural children and those whose mothers reported non-institutional delivery. However, the adjusted prevalence ratios did not show statistically significant associations with either residence or place of delivery. In the 2019 Mini Demographic and Health Survey, Muchie and colleagues did report adjusted associations with rural residence and noninstitutional delivery [3]. Maternal education, household financial situation and sanitation also showed no clear overall adjusted association in the present study. Earlier Ethiopian research on full vaccination found inequalities by wealth and maternal education [16]. Differences in vaccination outcomes, populations and covariate adjustment may contribute to these findings. Full vaccination measures completion of several doses, so those findings do not directly contradict results for first-dose nonreceipt. The absence of a clear adjusted association here does not establish that these characteristics are unrelated to vaccination access.

The overall association with walking time to the nearest health facility was not statistically significant (p = 0.052), although category-specific estimates were higher for longer walking times. Okwaraji and colleagues found lower third-pentavalent-dose coverage among children living at least an hour from a health post in rural northwestern Ethiopia [30]. Their study included children aged 12–59 months and used geographically estimated travel time, whereas the present survey used reported walking time and examined first-dose nonreceipt. The present estimates suggest a possible access barrier, although the overall association was not statistically conclusive.

Self-reported local armed conflict was associated with lower zero-dose prevalence (aPR: 0.70, 95% CI: 0.50–0.99), and children whose mothers decided alone on child healthcare had lower prevalence than those with joint decisions (aPR: 0.64, 95% CI: 0.44–0.94). The timing and intensity of reported conflict in relation to vaccination were not established. Household decision-making responses did not identify who arranged or provided vaccination. Qualitative research in underserved Ethiopian communities identified gender norms and limited support for caregivers as barriers to childhood vaccination [25]. The decision-making association may therefore reflect household circumstances not fully captured by the response categories. Both inverse associations may be affected by residual confounding and should not be interpreted as protective effects.

The associations of ANC attendance and participation in decisions about child healthcare with zero-dose status varied across sampled communities with adequate local data. Endehabtu and colleagues also reported geographic variation in associations involving ANC and vaccination perceptions [4]. Together, these findings suggest that national average associations may not describe every locality. The local maps can help guide assessment of maternal healthcare use and household decision-making in the identified communities.

The local analyses identified concentrations of high zero-dose prevalence within the broader regional pattern. Four communities were classified as high–high clusters and nine as hotspots in Somali; the primary scan cluster extended across parts of Somali, Oromia and Sidama. Nigerian surveys have also mapped zero-dose concentrations while examining first-dose status separately from other vaccination outcomes [31,32]. In Zambia, geospatial analysis found differences in zero-dose prevalence within and between health-facility catchment areas [33]. In Kikwit, Democratic Republic of the Congo, Mutwadi and colleagues found scan clusters spanning several health areas and associations with distance and absence of community-health-worker visits [34]. Although their city sample differs from a national survey, the findings illustrate how local concentrations can cross administrative borders. Coordination between neighbouring service areas may therefore help reach children in the identified clusters.

The predicted surface showed higher prevalence in parts of eastern and south-central Ethiopia, consistent with the clusters identified in sampled communities. Spatial cross-validation showed a modest reduction in prediction error compared with predictions based on the pooled prevalence in the training sample. Other spatial vaccination studies have demonstrated the value of detailed maps for identifying gaps while emphasizing the importance of survey design, geographic scale and prediction uncertainty [35–40]. The 10-km predictions can guide local assessment, although their accuracy is limited where sampled communities are sparse.

The 2024–25 Ethiopia Demographic and Health Survey reported first-pentavalent-dose receipt in 70.6% of children aged 12–23 months, equivalent to 29.4% without the dose, based on vaccination cards or maternal reports [41]. The present study found first-dose nonreceipt of 20.6% among children aged 12–35 months. Differences in age eligibility and survey methods limit direct comparison. Nevertheless, both surveys indicate a substantial unmet need for first-dose vaccination beyond infancy.

The mapped concentrations support prioritizing first-dose vaccination services in the identified communities. The primary scan zone crossed Somali, Oromia and Sidama, suggesting that reaching missed children may require coordination across neighbouring service areas. Reliable outreach and clear information for caregivers are relevant priorities, consistent with barriers reported in underserved Ethiopian settings [25]. Later-dose dropout, documented separately in these settings [42], also requires continued contact after children enter the schedule. Research on the immunization cascade across low- and middle-income countries also documents dropout after the first dose [43]. Identifying zero-dose children should therefore be accompanied by follow-up to complete the recommended doses.

## Strengths and limitations

This study drew on a national survey covering Ethiopia’s regional states and chartered cities, accounted for complex sampling in prevalence estimates and adjusted association analyses, and examined geographic clustering using several spatial methods.

This study finding might present with limitations such as a reporting bias as zero-dose status was assessed from vaccination records and caregiver reports; incomplete records and recall errors may affect classification. Several regional estimates were imprecise and some EAs included few eligible children. Maternal health-service information referred to the woman’s most recent eligible pregnancy and may not relate to the pregnancy of the child included in this analysis. The predicted values depend on the distribution of sampled EAs, which limits local interpretation, as discussed in other spatial vaccination studies. The cross-sectional design and correlated household and service factors preclude causal attribution.

## Conclusions

Approximately one in five children aged 12–35 months had not received the first pentavalent dose. ANC utilization, drinking-water sources, and caregivers’ perceived importance of vaccination were associated with higher zero-dose prevalence. High prevalence clustered among sampled communities in eastern and south-central Ethiopia. Immunization programmes should prioritize identifying and vaccinating missed children in these communities, including those aged 24–35 months, through age-appropriate catch-up services. Strengthening links between maternal healthcare and immunization, addressing caregiver concerns, and reviewing the regularity and reach of outreach services are relevant programme priorities. Local planning should combine the geographic findings with current service and household information.

## Data Availability

The data will be made available by Project HOPE upon reasonable request to the corresponding author.

## Declarations

### Ethical considerations

The survey protocol was reviewed and approved by the Ethiopian Public Health Institute Institutional Review Board (Protocol No. EPHI-IRB-567-2024). Administrative permissions were obtained from the Ministry of Health and relevant regional health bureaus. Informed consent was obtained from participants before data collection. De-identified survey data were used for this analysis.

### Author contributions

Mulugeta Geremew: Conceptualization, methodology, investigation, formal analysis, software, visualization, writing – original draft. Fisseha Shiferie: Conceptualization, methodology, investigation, supervision, writing – review and editing. Asnake Worku, Abyot Asres, Ayalew Astatkie, Tigist Shumet, Dawit A. Tsegaye, Gashaw Andargie, and Samson Gebremedhin: Conceptualization, methodology, investigation, writing – review and editing.

## Acknowledgements

The authors thank the survey participants, field teams, the Ethiopian Ministry of Health, Ethiopian Public Health Institute and Pathways for Ethiopia survey team.

## AI use declaration

OpenAI was used to edit the manuscript for grammar, spelling, and language errors. It was not used to perform the statistical analyses.

## Supporting information

S1 Table. Detailed survey-weighted regional estimates.

S2 Table. Local Moran and Getis–Ord Gi* categories after false-discovery-rate adjustment.

S3 Table. Region comparisons in the adjusted factor model.

S4 Table. Evidence for local coefficient mapping.

